# A single dose of cannabidiol modulates the coupling between hippocampal glutamate and learning-related prefrontal activation in individuals at Clinical High Risk of Psychosis

**DOI:** 10.1101/2024.11.06.24316831

**Authors:** Yiling Shi, Cathy Davies, Robin Wilson, Elizabeth Appiah-Kusi, David J. Lythgoe, Gemma Modinos, Sagnik Bhattacharyya

## Abstract

**Background:** Cannabidiol (CBD) is being studied as a potential intervention for the people at clinical high risk for psychosis (CHR), though the mechanisms underlying its effects are not fully understood. Previous studies indicate that a single dose of CBD can normalize alterations in memory-related brain activation and modulate hippocampal glutamate levels in the early stages of psychosis. This study aimed to examine the acute effects of CBD on the coupling between hippocampal glutamate levels and brain activation during verbal memory in individuals at CHR.

**Methods:** 33 participants at CHR participants were randomly assigned to receive either a single dose of 600 mg CBD (CHR-CBD) or an identical placebo capsule (CHR-PLB). 19 age-matched healthy controls (HC) received no study drugs. Participants underwent MRI scanning while performing a verbal learning task, and proton magnetic resonance spectroscopy to measure hippocampal glutamate levels. Group x hippocampal glutamate x brain activation interactions were tested.

**Results:** CHR-PLB showed positive correlation between hippocampal glutamate levels and dorsolateral prefrontal cortex (dlPFC) (Pcorr. = 0.0039) activation compared to HC during both verbal encoding and recall. Under a single dose of CBD, the glutamate-dlPFC activation coupling was negative and significantly different compared to placebo in CHR individuals (Pcorr. = 0.0001). The reversed correlation in CBD group also observed in the parahippocampal gyrus (Pcorr. = 0.0022) and amygdala (Pcorr. = 0.0019).

**Conclusions:** These findings suggest that the mechanism underlying the effect of CBD in CHR may involve reversing of altered coupling between hippocampal glutamate levels and prefrontal and mediotemporal activation.

## Introduction

The onset of psychotic disorders is often preceded by attenuated symptoms of psychosis, anxiety, emotional distress and a decline in function, collectively referred to as the clinical high-risk (CHR) state for psychosis (Yung et al., 2005). About a quarter individuals at CHR may develop a frank psychotic disorder within 3 years (Salazar de Pablo et al., 2021). There are currently no licensed pharmacological preventive treatments for this population and only modest evidence of the efficacy of available treatments (Davies et al., 2018a, 2018b; Devoe et al., 2020; Fusar-Poli et al., 2019). Novel therapeutics are therefore needed. Cannabidiol (CBD), the non-intoxicating component of cannabis, has demonstrated antipsychotic potential (Leweke et al., 2012; McGuire et al., 2017) and is well tolerated, thus holding promise as a potential intervention for the CHR state (Bhattacharyya et al., 2024; Davies & Bhattacharyya, 2019). However, the neurobiological mechanisms underlying the effects of CBD in CHR individuals remains unclear.

Evidence from preclinical and human neuroimaging studies indicates a central role for hippocampal glutamatergic dysfunction in the onset of psychosis, by driving hyperactivity in a hippocampal-midbrain-striatal circuit (Daniel J & Anthony A, 2011; Grace & Gomes, 2019; Lieberman et al., 2018; Schobel et al., 2013). This has been consistent with evidence of altered hippocampal glutamatergic concentrations (Kraguljac et al., 2013), increased hippocampal and striatal perfusion (Allen et al., 2016; Davies et al., 2023; Kraguljac et al., 2013; Schobel et al., 2009; Talati et al., 2015) and elevated striatal dopamine function (Egerton et al., 2013; Howes et al., 2011) across different stages of the psychosis continuum. Hippocampal glutamatergic dysfunction has been hypothesised in turn to arise as a result of dysfunction in the corticolimbic circuit from the prefrontal cortex to the hippocampus (Gomes & Grace, 2016; Lewis & Hashimoto, 2007), particularly affecting GABAergic interneurons (Smucny et al., 2022). Prefrontal GABA concentrations were found to be associated with increased resting hippocampal perfusion in CHR individuals who subsequently developed psychosis (Modinos et al., 2018), consistent with preclinical evidence linking prefrontal and hippocampal dysfunction and suggesting potential mechanisms underlying cognitive dysfunction in schizophrenia (Abbas et al., 2018; Dienel et al., 2023; Santos-Silva et al., 2024; Smucny et al., 2022).

Prior research examining interactions between neurochemistry and brain activation in psychosis showed positive correlations between hippocampal glutamate levels and memory-related prefrontal (Hutcheson et al., 2012) and medial temporal (Valli et al., 2011) activation in healthy controls, which were not present in individuals with psychosis (Hutcheson et al., 2012) or at CHR for psychosis (Valli et al., 2011). We have recently demonstrated (1) an altered relationship between hippocampal glutamate levels and striatal resting perfusion in individuals at CHR (Davies et al., 2023a), and (2) increased memory-related hippocampal-striatal connectivity in those with established psychosis (O’Neill, Wilson, et al., 2021). Notably, a single dose of CBD partially normalised the latter alterations (O’Neill, Wilson, et al., 2021) as well as hippocampal glutamate levels (O’Neill, Annibale, et al., 2021). However, whether a single dose of CBD can modulate the relationship between hippocampal glutamate levels and brain activation remains unclear.

To address this question, we investigated the coupling between hippocampal glutamate levels, indexed by proton magnetic resonance spectroscopy (1H-MRS), and brain activation as measured using functional magnetic resonance imaging (fMRI) during a verbal paired-associates learning task - a key cognitive domain shown to be particularly impaired people at CHR (Catalan et al., 2021). A region of interest (ROI) approach was used focusing on the striatum, medial temporal and prefrontal cortical regions – regions previously shown to be modulated during this task in healthy controls (Bhattacharyya et al., 2009), altered in psychosis (O’Neill, Annibale, et al., 2021), and normalised by CBD (Bhattacharyya et al., 2018; O’Neill, Wilson, et al., 2021). Three participant groups were examined: CHR individuals randomised to a single oral dose (600mg) of CBD (CHR-CBD) or placebo (CHR-PLB) and healthy controls.

We hypothesised that (1) the coupling between hippocampal glutamate levels and fMRI activation would be altered in CHR-PLB compared to healthy controls, and (2) a single dose of CBD would modulate this coupling in CHR individuals, bringing it closer to that seen in healthy controls. Specifically, we expected a positive correlation between hippocampal glutamate levels and activation in the prefrontal and medial temporal regions (Hutcheson et al., 2012; Valli et al., 2011), and a negative correlation in the striatum for healthy controls (Davies et al., 2023a). Additionally, we predicted that CBD (in a separate CHR group) would reverse the direction of the correlations seen in the CHR-PLB group, aligning them with those observed in healthy controls.

## Methods

### Participants

The analysis of stand-alone fMRI and ^1^H-MRS data included in this study have been separately reported in previous publications. (Bhattacharyya et al., 2018; Davies et al., 2023a). Briefly, 33 antipsychotic-naive CHR participants (aged 18-35), were recruited from early intervention services from the South London and Maudsley NHS Foundation Trust. 19 healthy controls, matched by age within ±3 years, were recruited through local advertisement. Participants with a history of psychotic or manic episodes, neurological disorders, substance dependence, IQ less than 70, and those who could not undergo MRI or be treated with CBD were excluded. Baseline levels of psychopathology were measured using the Comprehensive Assessment of At-Risk Mental States (CAARMS) (Yung et al., 2005) and the State-Trait Anxiety Inventory-State Subscale (STAI-S) (Spielberger et al., 1971). The study was approved by the National Research Ethics Service Committee of London Camberwell St Giles, and all participants provided written informed consent.

### Design

The study employed a double-blind, parallel-group, placebo-controlled design, in which CHR participants were randomly assigned to receive either a single dose of CBD (600mg; THC-Pharm) or placebo capsule. Detailed experiment procedures can be found in the Supplementary Material. This CBD dose has previously been shown to be effective in established psychosis (Leweke et al, 2012). Three hours after taking the CBD or placebo capsule, participants underwent MRI scanning, where hippocampal glutamate levels were measured using ^1^H-MRS followed by the fMRI verbal paired-associates (VPA) task (detailed below). The dosing-to-scanning interval was determined based on previous evidence showing that CBD levels in blood reach their peak around 3 hours following oral administration (Martin-Santos et al., 2012). The healthy control group was subjected to the same procedures as the CHR group but did not receive any study drugs. Before scanning, all participants were instructed to abstain from consuming cannabis for 96 hours, alcohol for at least 24 hours, nicotine for 6 hours and any other recreational drugs for two weeks before the study day. A urine sample was collected before scanning to screen for the use of illicit drugs.

### ^1^H-MRS acquisition and quantification

All MRI data was collected using a Signa HDx 3T MRI scanner (General Electric). A high-resolution inversion recovery image was also acquired. Full details of ^1^H-MRS acquisition parameters, preprocessing and analysis methods have been reported elsewhere (Davies et al., 2023a) and in Supplementary Material. In brief, we acquired ^1^H-MRS spectra using PRESS (Point RESolved Spectroscopy; TE = 30ms; TR = 3000ms; 96 averages) in the left hippocampus, following previously established methods (Egerton et al., 2014).

LCModel version 6.3-0A (Provencher, 1993) was used to analyse the acquired spectra, with a standard basis set of 16 metabolites (Supplementary Material) as detailed in the LCModel manual (http://s-provencher.com/pub/LCModel/manual/manual.pdf). Metabolite analyses were restricted to those meeting the quality control criteria of Cramér–Rao lower bounds ≤ 20% and signal to noise ratio ≥ 5. We corrected values of the water-scaled measure of glutamate for voxel tissue and cerebral spinal fluid (CSF) content (Egerton et al., 2014) (Supplementary Material). Assessment of spectral quality involved measuring both the signal-to-noise ratio and the linewidths, represented by the full width at half-maximum (FWHM).

### fMRI acquisition

The blood oxygen level-dependent (BOLD) hemodynamic response during the task was measured using a gradient echo sequence axially; 39 × 3mm slices; 0.3mm gap between slices running parallel to the intercomissural (AC-PC) plane (field of view 24 × 24 cm and matrix 64 × 64); 30ms echo time; 90° flip angle) and a compressed acquisition with a 2s repetition time and 3s silent period (effective TR = 5s).

### fMRI task

Participants were studied in a 13-minute fMRI experiment while performing a VPA task as previously described (Bhattacharyya et al., 2018). During three conditions: encoding, recall, and baseline, the 8 pairs of stimuli were presented visually in 40-second blocks with the conditions presented in the same order (encoding, recall and baseline) and repeated 4 times. Participants’ response accuracy was recorded in real time. During the encoding phase, word pairs were presented and to encourage encoding, participants were instructed to indicate aloud whether the two words were related or not. The same word pairs were presented four times during encoding (once per block) to aid in the learning of associations across repeated blocks. In the recall phase, one of the words from each of the previously presented pairs was displayed one at a time, and participants were asked to recall the word that it had been paired with. If they could not recall the missing word, they were told to say "pass". The baseline phase involved the participants viewing a pair of blank blue rectangles of the same size as those in the encoding/recall condition.

### fMRI processing

#### fMRI data preprocessing and subject-level analysis

fMRI data were analysed using XBAM v4.1 software (Brain Imaging Analysis Unit, Institute of Psychiatry, Kings College London, UK), which employs a nonparametric approach to minimize assumptions about the distribution of the data. The fMRI preprocessing and subjects-level analysis steps have been reported previously (Bhattacharyya et al., 2018a) and are detailed in the Supplementary Material. Briefly, before analysis, the images were corrected for head motion (Brammer et al., 1997; Bullmore et al., 1999), slice timing, and spatially smoothed with an 8.8mm full-width-at-half-maximum Gaussian filter (Thirion et al., 2007). The experimental design (encoding and recall condition) was convolved with 2 γ-variate functions. This model was then fit to the time series at each voxel using least-squares fitting. Following this, the sum of squares (SSQ) ratio statistic, which represents the ratio of the model component to the residual sum of squares, was estimated for the encoding and correct recall conditions relative to baseline. The SSQ ratio maps of each condition for each individual were then transformed into standard stereotactic (Talairach) space

#### Generation of ROI mask

The regions of interest (ROI) masks, including the bilateral medial temporal cortex (hippocampus and parahippocampal gyrus), striatum (caudate and putamen), and prefrontal cortex (inferior and middle frontal gyri), were generated using Talairach Atlas labels in XBAM (eFigure 2).

#### Pearson correlation calculation for each group (HC; CHR-PLB; CHR-CBD)

Using XBAM 4.1, we estimated the Pearson linear correlation coefficient (*r*) between hippocampal glutamate level and SSQ value for each contrast (encoding vs baseline, recall vs baseline) at each intracerebral voxel for each group separately within our ROI mask. The Pearson linear correlation coefficient (*R*) is defined as below:

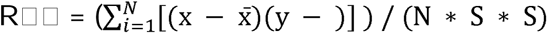

Where X is the estimated SSQ value and Y is the hippocampal glutamate level, x□ and □ the respective sample mean, S□ and S_y_ are the estimated standard deviations.

#### Correlation between hippocampal glutamate levels and brain activation in HC

To first understand the normative relationship between hippocampal glutamate and brain activation during the verbal learning task, we explored which brain regions showed a significant correlation with hippocampal glutamate levels in the HC group only using a method described before (Chantiluke et al., 2014). XBAM uses wavelet-based permutation techniques (E. Bullmore et al., 2001; E. T. Bullmore et al., 1999) to assess the statistical significance of the correlation coefficient. Under the null hypothesis, the estimated SSQ value for each voxel within the ROI mask and hippocampal glutamate levels were uncorrelated, meaning there is no underlying pairing structure between them. Consequently, the order of observations for the hippocampal glutamate level were independently permuted within each group. By permuting the hippocampal glutamate levels 1,000 times and calculating the correlation coefficient with the estimated SSQ value at each voxel within the ROI mask for these generated samples, we obtained the distribution of correlation coefficients under the null hypothesis that there is no relationship between hippocampal glutamate levels and the estimated SSQ values. We then determined the critical value of the correlation coefficient at any desired type 1 error level in the original non-permuted data with reference to the null distribution to evaluate the statistical significance of the observed correlation. We then extended the analysis to the 3D cluster level (E. T. Bullmore et al., 1999). This process was conducted for each contrast separately (encoding vs baseline; recall vs baseline).

#### Pairwise group differences in correlation analysis (HC vs CHR-PLB; CHR-PLB vs CHR-CBD)

To investigate pairwise group differences (HC vs CHR-PLB; and CHR-PLB vs CHR-CBD) in the association between hippocampal glutamate levels and brain activation (as indexed by the SSQ ratio statistic) in our regions of interest, we compared the correlation coefficients between each pair of groups (HC vs CHR-PLB, CHR-PLB vs CHR-CBD) for each of the two contrasts (encoding vs baseline; recall vs baseline) using a method described before (Murphy et al., 2014). For each group pair (HC vs CHR-PLB, CHR-PLB vs CHR-CBD) and for each contrast (encoding vs baseline; recall vs baseline), we independently estimated the average Pearson correlation coefficient between left hippocampal glutamate level and brain activation as indexed by the SSQ ratio statistic and calculated the group differences in correlation. To evaluate the significance of this difference, using a similar method as described above, we then generated a null distribution of differences in correlation within our regions of interest by randomly permuting subjects and hippocampal glutamate values between groups, thereby scrambling any between group differences. For each such permutation, we calculated correlation differences between the scrambled groups and combined the resulting values across all voxels to produce a null distribution of differences in correlation within our regions of interest. We then extended the analysis to the cluster level, setting the cluster probability under the null hypothesis at a level such that the expected number of type 1 error clusters was less than one. To control for multiple comparisons, for each contrast, we used a voxelwise statistical threshold of p < 0.05 and adjusted the clusterwise threshold to ensure less than one false-positive cluster over the whole map. For post-hoc visualisation purposes and to examine the direction of any associations, the average SSQ value of each significant cluster was extracted and correlated with hippocampal glutamate values, then plotted using the ggplot package in R v.4.2.3.

## RESULTS

### Demographic characteristics, ^1^H-MRS glutamate levels and verbal learning task performance

Demographic characteristics of the cohort, hippocampal glutamate levels and verbal learning task performance have been reported previously (Bhattacharyya et al., 2018; Davies et al., 2023a; Wilson et al., 2019) and are included here for completeness (Table 1). As reported before, there were no significant differences in demographic data except for fewer years of education in the CHR-PLB group relative to HC (Davies et al., 2023a). Compared to the HC group, individuals with CHR-placebo showed significantly lower levels of hippocampal glutamate (P= .015). Furthermore, a significant linear trend was detected across the groups, indicating that glutamate levels were highest in HC, lowest in the CHR-PLB group, and at an intermediate level in the CHR-CBD group (P = .031) (Davies et al., 2023). As reported before, there was no significant difference in recall performance accuracy rate (HC vs CHR-PLB; CHR-PLB vs CHR-CBD) among the three groups (Bhattacharyya et al., 2018). In the present study, fMRI data from three participants (one from each group) were excluded due to poor image quality. In total, data from 49 participants (CHR-PLB n=15, CHR-CBD n=16, HC n=18) was analysed.

**TABLE 1.**
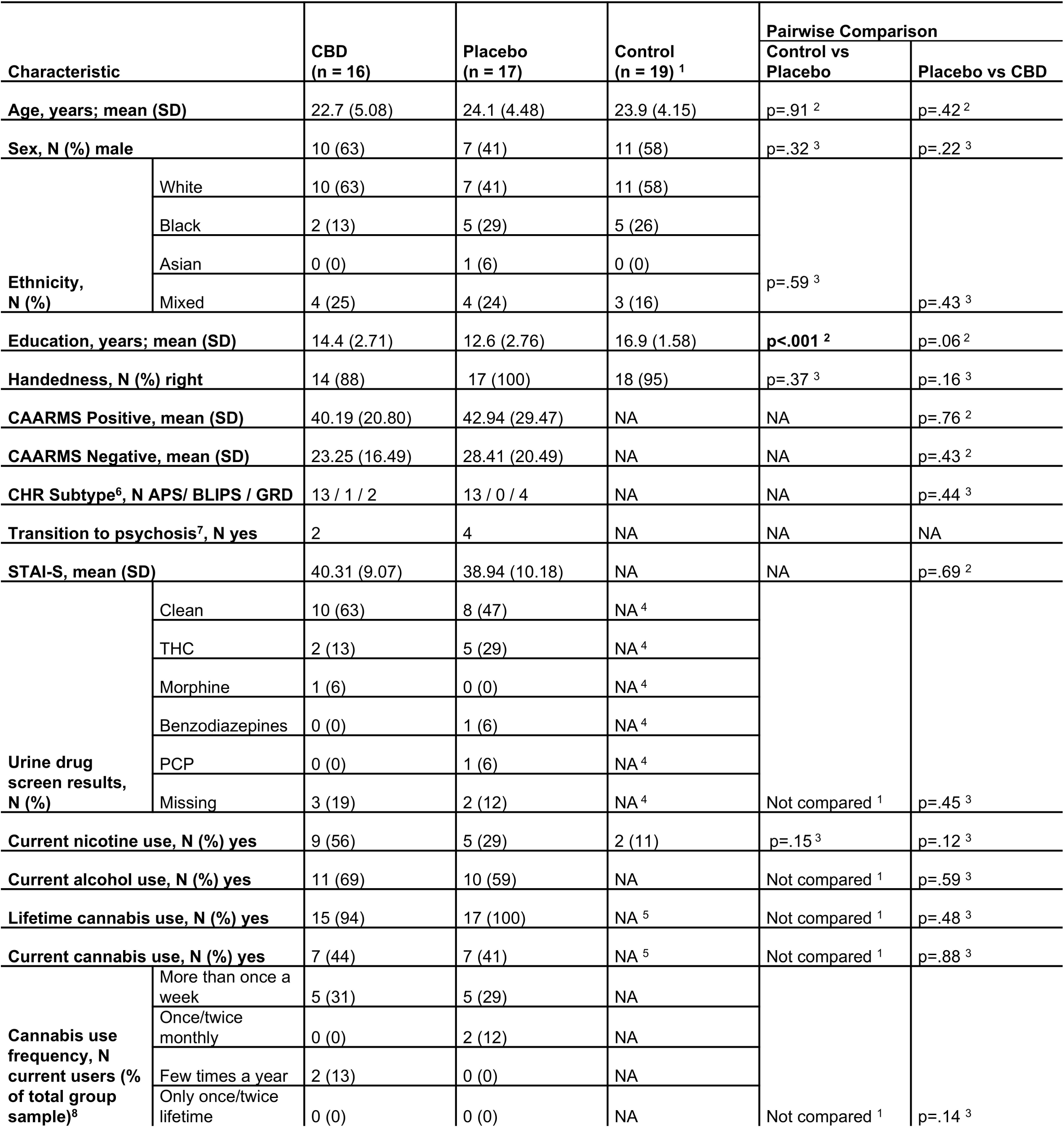
Sociodemographic and Clinical Characteristics at Baseline. *Abbreviations:* CAARMS, Comprehensive Assessment of At-Risk Mental States; CBD, cannabidiol; CHR, Clinical High Risk for Psychosis; N, number of subjects; NA, not applicable; PCP, phencyclidine; STAI-S, State-Trait Anxiety Inventory-State Subscale;wh THC, Δ9-tetrahydrocannabinol. ^1^Controls were selected to have minimal drug use and hence were not compared with CHR participants on these parameters; ^2^Independent t-test; ^3^Pearson chi-squared test; ^4^Controls tested negative on urine drug screen for all substances tested; ^5^Cannabis use less than 10 times lifetime (no current users); ^6^CAARMS subgroup: BLIPS brief limited intermittent psychotic symptoms, APS attenuated psychotic symptoms, GRD genetic risk and deterioration; ^7^Data on the later transition to psychosis was not systematically collected and thus these numbers should be interpreted with caution, particularly as the transition is a time-dependent outcome; ^8^The count data (N) represent the number of current cannabis users (of which there were 7 in each CHR group) who reported typical cannabis use at each given frequency. The p-value shown relates to this count data. Percentages reflect the % of the total group sample who are current cannabis users and typically using at each frequency.

### Correlation between hippocampal glutamate levels and brain activation in HC

In HC participants, during the encoding condition, hippocampal glutamate levels were significantly negatively correlated with activation in the left middle frontal gyrus (Pcorr. = 0.001) and right putamen (Pcorr. = 0.0029). During the recall condition, there was a significant negative correlation between hippocampal glutamate levels and activation in the left middle frontal gyrus (Pcorr. = 0.001) (Table 2 and eFigure 3).

**Table 2.** Interactions between the group, hippocampal glutamate and brain activation within ROIs during encoding and recall conditions.

| Region | Coordinates of Peak (TAL) |  |  | Cluster Size,<br>No. of<br>Voxels | Corrected P<br>Value<sup>a</sup> |
| --- | --- | --- | --- | --- | --- |
|  | X | Y | Z |  |  |
| Healthy Control |  |  |  |  |  |
| Verbal Encoding |  |  |  |  |  |
| L Middle Frontal Gyrus | -29 | 22 | 46 | 24 | 0.0001 |
| R Putamen | 22 | 7 | -10 | 10 | 0.0029 |
| Verbal Recall |  |  |  |  |  |
| L Middle Frontal Gyrus | -43 | 22 | 30 | 14 | 0.0010 |
| Healthy Control vs CHR-PLB |  |  |  |  |  |
| Verbal Encoding |  |  |  |  |  |
| L Middle frontal gyrus<br>(extending to precentral<br>gyrus) | -41 | 3 | 42 | 19 | 0.0039 |
| L Middle Frontal Gyrus | -47 | 33 | -7 | 4 | 0.0056 |
| Verbal Recall |  |  |  |  |  |
| R Inferior Frontal Gyrus | 43 | 30 | 7 | 15 | 0.0024 |
| L Middle Frontal Gyrus | -43 | 22 | 30 | 16 | 0.0017 |
| CHR-PLB vs CHR-CBD |  |  |  |  |  |
| Verbal Encoding |  |  |  |  |  |
| L Precentral Gyrus | -32 | 4 | 30 | 7 | 0.0008 |
| L Precentral Gyrus<br>(extending to middle<br>frontal gyrus) | -40 | 7 | 36 | 40 | 0.0001 |
| Verbal Recall |  |  |  |  |  |
| L Inferior Frontal Gyrus | -51 | 4 | 13 | 15 | 0.0008 |
| L Precentral Gyrus | -32 | 4 | 30 | 12 | 0.0023 |
| L Parahippocampal Gyrus | -22 | -43 | -9 | 12 | 0.0022 |
| L Amygdala | -25 | -7 | -16 | 9 | 0.0019 |

L: Left side of the brain; R: Right side of the brain

### Group differences in the relationship between hippocampal glutamate levels and brain activation

#### HC vs CHR-PLB

There was a significant difference between the HC and CHR-PLB groups in terms of the relationship between hippocampal glutamate levels and regional brain activation. Specifically, during the encoding condition, a positive relationship was observed between hippocampal glutamate and brain activation in a left middle frontal gyrus (ventrolateral prefrontal cortex) cluster in HC, while a negative relationship was observed in CHR-PLB individuals (Pcorr. = 0.0039) (Table 2; Figure 1A). This was reversed in an adjacent cluster located on the more ventral caudal side of the left middle frontal gyrus (dorsolateral prefrontal cortex), where there was a negative relationship in healthy controls and a positive relationship in CHR-PLB individuals (Pcorr. = 0.0056) (Table 2; Figure 1A). During the recall condition, there was a negative relationship between hippocampal glutamate and activation in the right inferior frontal gyrus (Pcorr. = 0.0024) and left middle frontal gyrus (Pcorr. = 0.0017) in HC, while there was a positive relationship in the CHR-PLB group (Table 2; Figure 1B).

**Figure 1:**
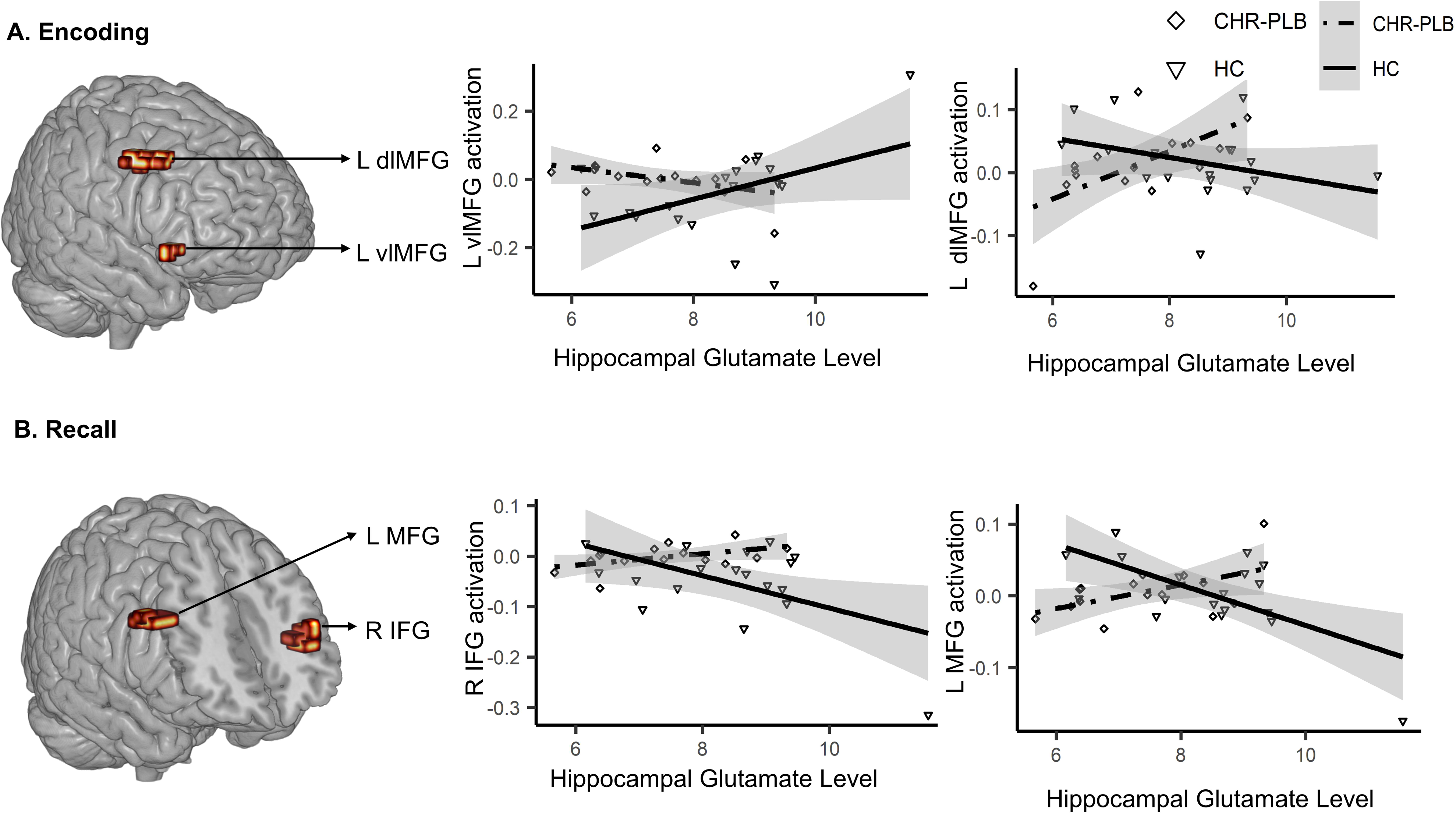
Clusters of brain activation (as indexed by the sum of squares ratio, SSQ) showing significant correlations with left hippocampal glutamate levels in the CHR-PLB vs HC comparison. The brain images depict significant clusters identified in the glutamate x BOLD x group interaction analysis, and the scatterplots show the relationship between hippocampal glutamate and the SSQ values extracted from each corresponding cluster. ***A. Encoding:*** The left scatter plot shows a cluster in the left ventrolateral middle frontal cortex (peak Talairach coordinates: X = −41, Y = 3, Z = 42; Cluster size = 19 voxels; p = 0.0039). The right scatter plot shows a cluster in the left dorsolateral middle frontal gyrus (peak Talairach coordinates: X = −47, Y = 33, Z = −7; Cluster size = 4 voxels; p = 0.0056). ***B. Recall:*** The left scatter plot shows a cluster in the right inferior frontal gyrus (R IFG) (peak Talairach coordinates: X = 43, Y = 30, Z = 7; Cluster size = 15 voxels; p = 0.0024). The right scatter plot displays another cluster in the left middle frontal gyrus (L MFG) (peak Talairach coordinates: X = −43, Y = 22, Z = 30; Cluster size = 16 voxels; p = 0.0017). This post-hoc analysis was used only for visualisation purposes and to determine the direction of the relationships between hippocampal glutamate and SSQ values as extracted from significant clusters. *Abbreviation: L – left side; R – right side*.

#### CHR-PLB vs CHR-CBD

On direct comparison of the CHR-PLB group with the CHR-CBD group, during the encoding condition, the CHR-PLB group exhibited a positive association between left hippocampal glutamate levels and brain activity in two adjacent clusters in the left precentral gyrus, one of them extending to the middle frontal gyrus, while where there was an inverse relationship in the CHR-CBD group in these clusters (Pcorr. = 0.0008; Pcorr. = 0.0001) (Table 2; Figure 2A). During the recall condition, there was a positive relationship between left hippocampal glutamate levels and activation in left inferior frontal gyrus (Pcorr. = 0.0008), precentral gyrus (Pcorr. = 0.0023), in the left parahippocampal gyrus (Pcorr. = 0.0022) and left amygdala (Pcorr. = 0.0019) in CHR-PLB individuals, while there was an inverse relationship in the CHR-CBD group in the same regions (Table 2; Figure 2B).

**Figure 2:**
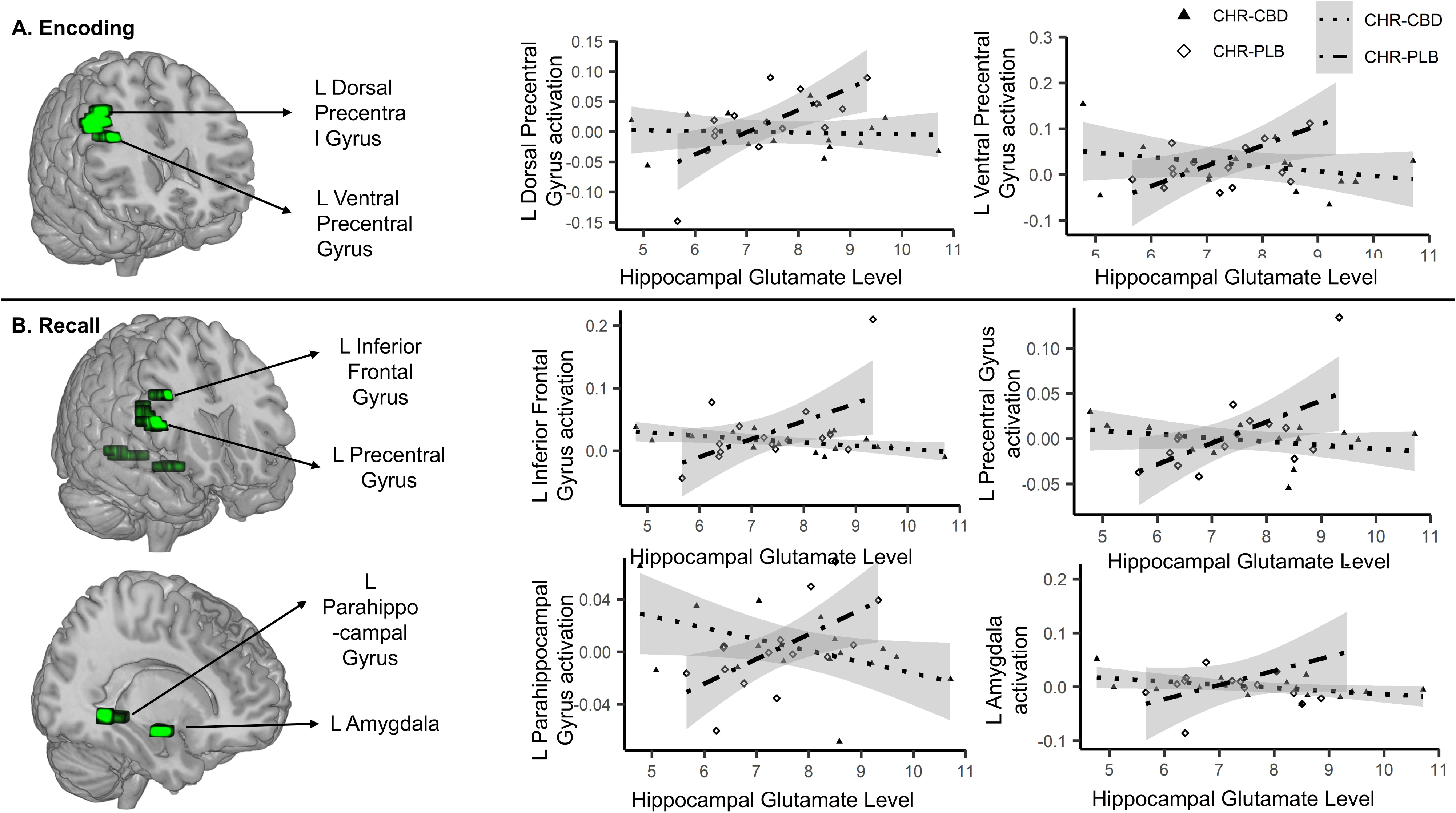
Clusters of brain activation (as indexed by the sum of squares ratio, SSQ) showing significant correlations with left hippocampal glutamate levels in the CHR-PLB vs CHR-CBD comparison. The brain images depict significant clusters identified in the glutamate x BOLD x group interaction analysis, and the scatterplots illustrate the relationship between hippocampal glutamate and the SSQ values extracted from each corresponding cluster. A. Encoding: The left scatter plot shows a cluster located in the left dorsal precentral gyrus (peak Talairach coordinates: X = −40, Y = 7, Z = 36; Cluster size = 40 voxels; p = 0.0001). The right scatter plot shows a cluster in the left ventral precentral gyrus (peak Talairach coordinates: X = −32, Y = 4, Z = 30; Cluster size = 7 voxels; p = 0.0008). B. Recall: The upper left scatter plot shows a cluster in the left inferior frontal gyrus (peak Talairach coordinates: X = −51, Y = 4, Z = 13; Cluster size = 15 voxels; p = 0.0008). The upper right scatter plot depicts a cluster in the left precentral gyrus (peak Talairach coordinates: X = −32, Y = 4, Z = 30; Cluster size = 12 voxels; p = 0.0023). The lower left scatter plot shows a cluster located in the left parahippocampal gyrus (peak Talairach coordinates: X = −22, Y = −43, Z = 9; Cluster size = 12 voxels; p = 0.0022). The lower right scatter plot displays a cluster in the left amygdala (peak Talairach coordinates: X = −25, Y = −7, Z = −16; Cluster size = 9 voxels; p = 0.0019). This post-hoc analysis was used only for visualisation purposes and to determine the direction of the relationships between hippocampal glutamate levels and SSQ values as extracted from significant clusters. *Abbreviation: L – left side*.

Overlaying the different group comparisons (HC vs CHR-PLB and CHR-PLB vs CHR-CBD) for encoding and recall conditions showed there was spatial overlap of the cluster of differential association with glutamate levels for the HC vs CHR-PLB comparison, with the cluster of differential association with glutamate levels for the CHR-PLB vs CHR-CBD comparison, for both encoding and recall conditions (Figure 3).

**Figure 3.**
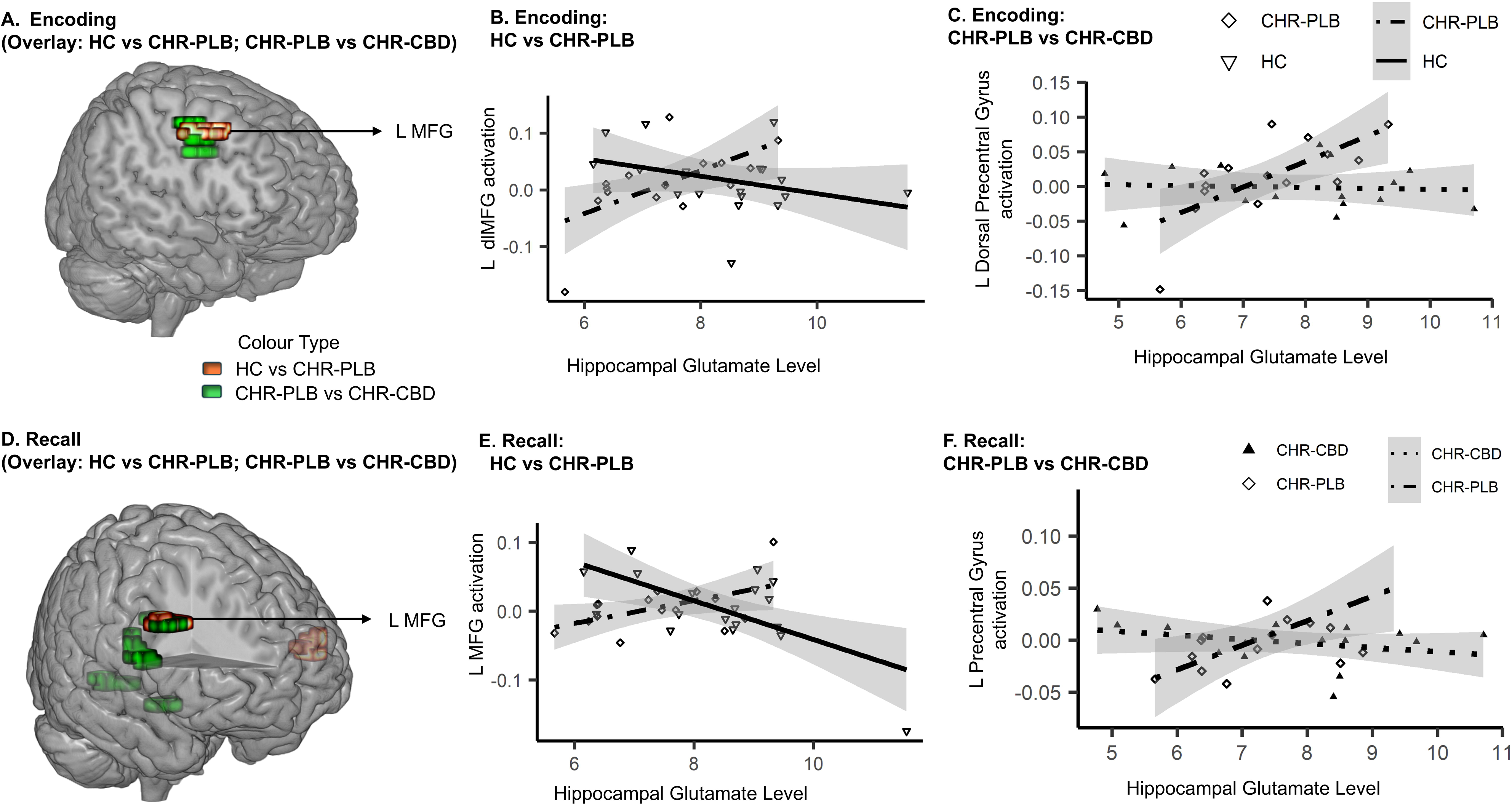
This figure displays an integrative visualization showing the overlap of clusters of differential activation across the two task conditions: Encoding (Scatter plots A and Line plots in B, C) and Recall (Scatter plots D and Line plots in E, F). The overlays and scatterplots highlight key areas of differential activation between-group contrasts—HC vs CHR-PLB and CHR-PLB vs CHR-CBD—focusing on the left dorsolateral middle frontal gyrus (L MFG). In Scatter plots A and D, the green clusters represent CHR-PLB vs CHR-CBD, while the orange and red clusters represent HC vs CHR-PLB during Encoding (A) and Recall (D). *Line plot B* corresponds to the orange cluster in Scatter plot A (dorsolateral prefrontal gyrus, peak talairach coordinate: X = −41, Y = 3, Z = 42; 19 voxels; p = 0.0039). *Line plot C* aligns with the green cluster in Scatter plot A (dorsolateral prefrontal gyrus, peak talairach coordinate: X = −43, Y = 22, Z = 30; 40 voxels; p = 0.0017). *Line plot E* matches the orange cluster in Scatter plot D (prefrontal gyrus, peak talairach coordinate: X = −40, Y = 7, Z = 36; 40 voxels; p = 0.0001). *Line plot F* matches the green cluster in Scatter plot B (Left Inferior Frontal Gyrus, peak talairach coordinate: X = −51, Y = 4, Z = 13; 15 voxels; p = 0.0008). *Abbreviation: L-left side*.

## Discussion

This study is, to the best of our knowledge, the first report on the effect of CBD on the coupling between hippocampal glutamate levels and brain fMRI activation during verbal learning, in CHR individuals compared to placebo and to a sample of HC (Bhattacharyya et al., 2018). Our key results are related to a pattern of correlation between hippocampal glutamate levels and learning-related brain activation that partially overlapped across the main comparisons of interest (HC vs CHR-PLB and CHR-CBD vs CHR-PLB) in this study. Specifically, relative to HC, we found that CHR participants under placebo showed a significantly altered correlation between hippocampal glutamate levels and activation in the dorsolateral prefrontal cortex during the encoding condition. In HC, there was a negative correlation between hippocampal glutamate and encoding-related engagement of the dorsolateral prefrontal cortex, while in the CHR-PLB group there was a positive correlation between hippocampal glutamate and activation in this region. Notably, we also observed that, compared to CHR-PLB participants, a single dose of CBD modulated the correlation between hippocampal glutamate and dorsolateral prefrontal activation in a partially overlapping region of the dorsolateral prefrontal cortex in a manner similar to the relationship observed in HC. As observed in the HC vs CHR-PLB contrast, in the comparison between CHR-PLB and CHR-CBD, we found a positive correlation between left hippocampal glutamate levels and encoding-related dorsolateral prefrontal activation in the CHR-PLB group, while there was an opposite relationship in the CHR-CBD group. We observed a similar overlapping pattern of correlation when considering the main comparisons of interest (HC vs CHR-PLB and CHR-CBD vs CHR-PLB) during the recall condition, such that it was also in a partially overlapping part of the dorsolateral prefrontal cluster observed in comparisons during the encoding condition.

Our results confirmed our initial prediction that, in individuals at CHR for psychosis, the relationship between hippocampal glutamate levels and verbal learning-related activation in the prefrontal cortex is opposite to that observed in HC(HC: negative; CHR-PLB: positive). Although the direction of correlation is the opposite of what we had predicted (HC: positive; CHR-PLB: negative), our results still suggest an abnormal coupling between hippocampal glutamate levels and prefrontal function associated with the risk of psychosis and are broadly consistent with other evidence from studies in people with established psychosis (Hutcheson et al., 2012; Nelson et al., 2022). However, the direction of the relationship differs between different studies, perhaps reflecting differences in imaging parameters (such as voxel positioning), specific task demands and patient cohort/ stage of illness. Our results find evidence partially in support of our second hypothesis that group differences (CHR-PLB vs CHR-CBD) in the correlation between hippocampal glutamate and activation in our chosen ROIs would overlap with regions where this relationship is different between HC and CHR-PLB, as we observed this for dorsolateral prefrontal cortex, but not for medial temporal and striatal ROIs. However, since there were no significant differences in task performance among the three groups, and we were unable to test the correlation between task performance and the coupling of glutamate and brain activation, the links between changes in neuronal activity and functional impairments or symptom severity remain unclear. Another possible explanation for our findings is that the reversal in the direction of the relationship between hippocampal glutamate levels and prefrontal activation may suggest that individuals at CHR engage the pathway between hippocampal glutamate and prefrontal cortex differently to complete the memory task at the same level as HC. In other words, this change might represent a compensatory mechanism in CHR individuals. In this context, the effect of CBD could be understood as alleviating the need for such compensation, allowing CHR individuals to disengage from this altered pathway when CBD is administered. Future studies could focus on exploring the relationship between task performance/ severity of symptoms and the correlation between hippocampal glutamate levels and dlPFC activation.

Our finding that the relationship between hippocampal glutamate levels and prefrontal cortex activation is altered in CHR individuals is consistent with the preclinical evidence suggesting that hippocampal dysfunction putatively driven by prefrontal cortex disruption (Gomes & Grace, 2016; Lewis & Hashimoto, 2007) is central to psychosis vulnerability (Daniel J & Anthony A, 2011; Grace & Gomes, 2019; Lieberman et al., 2018; Schobel et al., 2013). Results presented here suggest that CBD may mitigate this dysfunction by normalizing the abnormal coupling between hippocampal glutamate levels and prefrontal cortex activation in people at CHR for psychosis. However, it is worth noting that as we employed a parallel group rather than a within-subject crossover design, we cannot be certain that CBD normalised the abnormal coupling in the same individuals in whom it was abnormal. Previous evidence indicates that medication effects and illness duration can influence the relationship between the glutamatergic system and neural activity (Zahid et al., 2023). However, since our antipsychotic-naïve subjects were early in the psychosis continuum, the altered correlations observed between hippocampal glutamate and task-related brain activation (HC vs. CHR-PLB and CHR-PLB vs. CHR-CBD) are unlikely to be confounded by illness duration or treatment effects.

Additional regions where CBD modulated the correlation between hippocampal glutamate and brain activation involved the left amygdala and parahippocampal gyrus. This is in line with previous studies showing that a single dose of CBD augments parahippocampal activation in healthy volunteers while they performed a same learning task as used in the present study (Bhattacharyya et al., 2010). Evidence suggests that CBD may modulate the level of Glx (collective levels of glutamate and glutamine) in individuals with autism spectrum disorders (Pretzsch et al., 2019), prefrontal and medial temporal activation during verbal memory processing (Bhattacharyya et al., 2018) and increased hippocampal glutamate levels in people with established psychosis (O’Neill, Annibale, et al., 2021) and CHR (trend level) (Davies et al., 2023a). Together with our findings, this implies that CBD’s antipsychotic effects (Bhattacharyya et al., 2024; Leweke et al., 2012, 2021; McGuire et al., 2017) may involve the normalisation of altered coupling between hippocampal glutamate levels and prefrontal and mediotemporal activation.

The molecular mechanisms underlying the effects of CBD are thought to involve multiple pathways and remain incompletely understood (Bih et al., 2015). A recent preclinical study suggested that CBD may specifically modulate excitatory-inhibitory (E/I) balance in the hippocampus by blocking GPR55-dependent pro-excitatory and anti-inhibitory effects (Rosenberg et al., 2023). E/I balance is proposed to be crucial for synchronizing signalling across diverse cortical regions (Uhlhaas, 2013) and has been strongly implicated in psychosis (Knight et al., 2022; Lányi et al., 2024; Uhlhaas & Singer, 2010). Our findings that CBD modulates the coupling between hippocampal glutamate levels and prefrontal, parahippocampal, and amygdala activity, provide indirect support for the hypothesis that CBD influences hippocampal glutamate levels, maintains a balanced E/I ratio, and preserves synchronized neural circuit function. These findings extend current understanding by suggesting that CBD’s modulation of glutamate activity may be a key mechanism through which it exerts therapeutic effects in disorders characterized by disrupted E/I balance, such as psychosis. Future research should aim to bridge the gap between preclinical findings and human studies by integrating preclinical techniques with neuroimaging methods such as fMRI and PET.

### Limitations

Given that our study was designed and powered for examining differences in neurophysiological markers rather than behavioural or clinical outcomes, our sample size was relatively modest. Future studies with larger sample sizes are now needed to identify the effects of CBD on task performance and symptoms and link these changes to brain function. Second, the parallel-group design means that we cannot definitively attribute the observed CHR-PLB vs CHR-CBD differences in glutamate-activation correlations solely to CBD administration, since inter-subject variability may contribute to results. To more precisely isolate the effects of CBD on CHR individuals and explore the underlying mechanisms, future research should consider a within-subject longitudinal design that includes repeated dosing of CBD.

## Conclusion

To the best of our knowledge, this is the first study to demonstrate that a single dose of CBD may partially attenuate the altered coupling between hippocampal glutamate levels and activation in the prefrontal cortex, amygdala, and parahippocampal regions in individuals at CHR. Future studies need to investigate this effect with repeated doses of CBD, larger cohorts of CHR individuals, and the use of preclinical strategies to further elucidate the underlying molecular mechanisms.

## Supporting information

Supplemental Materia

## Data Availability

All data produced in the present study are available upon reasonable request to the authors

## Acknowledgements

The authors thank the study volunteers, the radiographers at the Centre for Neuroimaging Sciences, King’s College London, who carried out the MRI scans.

## Ethical standards

The authors assert that all procedures contributing to this work comply with guidance from the National Research Ethics Service Committee of London Camberwell St Giles institutional on human experimentation.

## Author Contributions

Substantial contributions to conception and design (SB, DJL), acquisition of data (RW, EAK), analysis and/or interpretation of data (YL, SB), drafting the article (YL, SB, GM, CD) or revising it critically for important intellectual content (all authors), study supervision (SB), final approval of the version to be published (all authors).

## REFERENCES

Abbas, A. I., Sundiang, M. J. M., Henoch, B., Morton, M. P., Bolkan, S. S., Park, A. J., Harris, A. Z., Kellendonk, C., & Gordon, J. A. (2018). Somatostatin Interneurons Facilitate Hippocampal-Prefrontal Synchrony and Prefrontal Spatial Encoding. Neuron, 100(4), 926–939.e3. 10.1016/j.neuron.2018.09.029

Allen, P., Chaddock, C. A., Egerton, A., Howes, O. D., Bonoldi, I., Zelaya, F., Bhattacharyya, S., Murray, R., & McGuire, P. (2016). Resting Hyperperfusion of the Hippocampus, Midbrain, and Basal Ganglia in People at High Risk for Psychosis. American Journal of Psychiatry, 173(4), 392–399. 10.1176/appi.ajp.2015.15040485

Bhattacharyya, S., Appiah-Kusi E, Wilson R, & McGuire P. (2024). Effects of cannabidiol on symptoms in people at clinical high risk for psychosis. World Psychiatry.

Bhattacharyya, S., Fusar-Poli, P., Borgwardt, S., Martin-Santos, R., Nosarti, C., O’Carroll, C., Allen, P., Seal, M. L., Fletcher, P. C., Crippa, J. A., Giampietro, V., Mechelli, A., Atakan, Z., & McGuire, P. (2009). Modulation of Mediotemporal and Ventrostriatal Function in Humans by Δ9-Tetrahydrocannabinol: A Neural Basis for the Effects of Cannabis sativa on Learning and Psychosis. Archives of General Psychiatry, 66(4), 442–451. 10.1001/archgenpsychiatry.2009.17

Bhattacharyya, S., Morrison, P. D., Fusar-Poli, P., Martin-Santos, R., Borgwardt, S., Winton-Brown, T., Nosarti, C., O’ Carroll, C. M., Seal, M., Allen, P., Mehta, M. A., Stone, J. M., Tunstall, N., Giampietro, V., Kapur, S., Murray, R. M., Zuardi, A. W., Crippa, J. A., Atakan, Z., & McGuire, P. K. (2010). Opposite Effects of Δ-9-Tetrahydrocannabinol and Cannabidiol on Human Brain Function and Psychopathology. Neuropsychopharmacology, 35(3), 764–774. 10.1038/npp.2009.184

Bhattacharyya, S., Wilson, R., Appiah-Kusi, E., O’Neill, A., Brammer, M., Perez, J., Murray, R., Allen, P., Bossong, M. G., & McGuire, P. (2018). Effect of Cannabidiol on Medial Temporal, Midbrain, and Striatal Dysfunction in People at Clinical High Risk of Psychosis: A Randomized Clinical Trial. JAMA Psychiatry, 75(11), 1107–1117. 10.1001/jamapsychiatry.2018.2309

Bih, C. I., Chen, T., Nunn, A. V. W., Bazelot, M., Dallas, M., & Whalley, B. J. (2015). Molecular Targets of Cannabidiol in Neurological Disorders. Neurotherapeutics, 12(4), 699–730. 10.1007/s13311-015-0377-3

Bullmore, E., Long, C., Suckling, J., Fadili, J., Calvert, G., Zelaya, F., Carpenter, T. A., & Brammer, M. (2001). Colored noise and computational inference in neurophysiological (fMRI) time series analysis: Resampling methods in time and wavelet domains. Human Brain Mapping, 12(2), 61–78. 10.1002/1097-0193(200102)12:2<61::AID-HBM1004>3.0.CO;2-W

Bullmore, E. T., Suckling, J., Overmeyer, S., Rabe-Hesketh, S., Taylor, E., & Brammer, M. J. (1999). Global, voxel, and cluster tests, by theory and permutation, for a difference between two groups of structural MR images of the brain. IEEE Transactions on Medical Imaging, 18(1), 32–42. IEEE Transactions on Medical Imaging. 10.1109/42.750253

Catalan, A., Salazar de Pablo, G., Aymerich, C., Damiani, S., Sordi, V., Radua, J., Oliver, D., McGuire, P., Giuliano, A. J., Stone, W. S., & Fusar-Poli, P. (2021). Neurocognitive Functioning in Individuals at Clinical High Risk for Psychosis: A Systematic Review and Meta-analysis. JAMA Psychiatry. 10.1001/jamapsychiatry.2021.1290

Chantiluke, K., Christakou, A., Murphy, C. M., Giampietro, V., Daly, E. M., Ecker, C., Brammer, M., Murphy, D. G., & Rubia, K. (2014). Disorder-specific functional abnormalities during temporal discounting in youth with Attention Deficit Hyperactivity Disorder (ADHD), Autism and comorbid ADHD and Autism. Psychiatry Research: Neuroimaging, 223(2), 113–120. 10.1016/j.pscychresns.2014.04.006

Daniel J, L., & Anthony A, G. (2011). Hippocampal dysregulation of dopamine system function and the pathophysiology of schizophrenia. Trends in Pharmacological Sciences, 32(9). 10.1016/j.tips.2011.05.001

Davies, C., & Bhattacharyya, S. (2019). Cannabidiol as a potential treatment for psychosis. Therapeutic Advances in Psychopharmacology. 10.1177/2045125319881916

Davies, C., Bossong, M. G., Martins, D., Wilson, R., Appiah-Kusi, E., Blest-Hopley, G., Allen, P., Zelaya, F., Lythgoe, D. J., Brammer, M., Perez, J., McGuire, P., & Bhattacharyya, S. (2023a). Hippocampal Glutamate, Resting Perfusion and the Effects of Cannabidiol in Psychosis Risk. Schizophrenia Bulletin Open, 4(1), sgad022. 10.1093/schizbullopen/sgad022

Davies, C., Bossong, M. G., Martins, D., Wilson, R., Appiah-Kusi, E., Blest-Hopley, G., Zelaya, F., Allen, P., Brammer, M., Perez, J., McGuire, P., & Bhattacharyya, S. (2023b). Increased hippocampal blood flow in people at clinical high risk for psychosis and effects of cannabidiol. Psychological Medicine, 1–11. 10.1017/S0033291723002775

Davies, C., Cipriani, A., Ioannidis, J. P. A., Radua, J., Stahl, D., Provenzani, U., McGuire, P., & Fusar-Poli, P. (2018b). Lack of evidence to favor specific preventive interventions in psychosis: A network meta-analysis. World Psychiatry: Official Journal of the World Psychiatric Association (WPA), 17(2), 196–209. 10.1002/wps.20526

Davies, C., Radua, J., Cipriani, A., Stahl, D., Provenzani, U., McGuire, P., & Fusar-Poli, P. (2018a). Frontiers | Efficacy and Acceptability of Interventions for Attenuated Positive Psychotic Symptoms in Individuals at Clinical High Risk of Psychosis: A Network Meta-Analysis. 10.3389/fpsyt.2018.00187

Devoe, D. J., Farris, M. S., Townes, P., & Addington, J. (2020). Interventions and Transition in Youth at Risk of Psychosis: A Systematic Review and Meta-Analyses. The Journal of Clinical Psychiatry, 81(3). 10.4088/JCP.17r12053

Dienel, S. J., Fish, K. N., & Lewis, D. A. (2023). The Nature of Prefrontal Cortical GABA Neuron Alterations in Schizophrenia: Markedly Lower Somatostatin and Parvalbumin Gene Expression without Missing Neurons. The American Journal of Psychiatry, 180(7), 495–507. 10.1176/appi.ajp.20220676

Egerton, A., Chaddock, C. A., Winton-Brown, T. T., Bloomfield, M. A. P., Bhattacharyya, S., Allen, P., McGuire, P. K., & Howes, O. D. (2013). Presynaptic Striatal Dopamine Dysfunction in People at Ultra-high Risk for Psychosis: Findings in a Second Cohort. Biological Psychiatry, 74(2), 106–112. 10.1016/j.biopsych.2012.11.017

Egerton, A., Stone, J. M., Chaddock, C. A., Barker, G. J., Bonoldi, I., Howard, R. M., Merritt, K., Allen, P., Howes, O. D., Murray, R. M., McLean, M. A., Lythgoe, D. J., O’Gorman, R. L., & McGuire, P. K. (2014). Relationship Between Brain Glutamate Levels and Clinical Outcome in Individuals at Ultra High Risk of Psychosis. Neuropsychopharmacology, 39(12), Article 12. 10.1038/npp.2014.143

Fusar-Poli, P., Davies, C., Solmi, M., Brondino, N., De Micheli, A., Kotlicka-Antczak, M., Shin, J. I., & Radua, J. (2019). Frontiers | Preventive Treatments for Psychosis: Umbrella Review (Just the Evidence). 10.3389/fpsyt.2019.00764

Gomes, F. V., & Grace, A. A. (2016). Prefrontal Cortex Dysfunction Increases Susceptibility to Schizophrenia-Like Changes Induced by Adolescent Stress Exposure. Schizophrenia Bulletin, sbw156. 10.1093/schbul/sbw156

Grace, A. A., & Gomes, F. V. (2019). The Circuitry of Dopamine System Regulation and its Disruption in Schizophrenia: Insights Into Treatment and Prevention. Schizophrenia Bulletin, 45(1), 148–157. 10.1093/schbul/sbx199

Howes, O., Bose, S., Turkheimer, F., Valli, I., Egerton, A., Stahl, D., Valmaggia, L., Allen, P., Murray, R., & McGuire, P. (2011). Progressive increase in striatal dopamine synthesis capacity as patients develop psychosis: A PET study. Molecular Psychiatry, 16(9), 885–886. 10.1038/mp.2011.20

Hutcheson, N. L., Reid, M. A., White, D. M., Kraguljac, N. V., Avsar, K. B., Bolding, M. S., Knowlton, R. C., den Hollander, J. A., & Lahti, A. C. (2012). Multimodal analysis of the hippocampus in schizophrenia using proton magnetic resonance spectroscopy and functional magnetic resonance imaging. Schizophrenia Research, 140(1–3), 136–142. 10.1016/j.schres.2012.06.039

Knight, S., McCutcheon, R., Dwir, D., Grace, A. A., O’Daly, O., McGuire, P., & Modinos, G. (2022). Hippocampal circuit dysfunction in psychosis. Translational Psychiatry, 12(1), Article 1. 10.1038/s41398-022-02115-5

Kraguljac, N. V., White, D. M., Reid, M. A., & Lahti, A. C. (2013). Increased Hippocampal Glutamate and Volumetric Deficits in Unmedicated Patients With Schizophrenia. JAMA Psychiatry, 70(12), 1294. 10.1001/jamapsychiatry.2013.2437

Lányi, O., Koleszár, B., Schulze Wenning, A., Balogh, D., Engh, M. A., Horváth, A. A., Fehérvari, P., Hegyi, P., Molnár, Z., Unoka, Z., & Csukly, G. (2024). Excitation/inhibition imbalance in schizophrenia: A meta-analysis of inhibitory and excitatory TMS-EMG paradigms. Schizophrenia, 10(1), 1–12. 10.1038/s41537-024-00476-y

Leweke, F. M., Piomelli, D., Pahlisch, F., Muhl, D., Gerth, C. W., Hoyer, C., Klosterkötter, J., Hellmich, M., & Koethe, D. (2012). Cannabidiol enhances anandamide signaling and alleviates psychotic symptoms of schizophrenia. Translational Psychiatry, 2(3), e94–e94. 10.1038/tp.2012.15

Leweke, F. M., Rohleder, C., Gerth, C. W., Hellmich, M., Pukrop, R., & Koethe, D. (2021). Cannabidiol and Amisulpride Improve Cognition in Acute Schizophrenia in an Explorative, Double-Blind, Active-Controlled, Randomized Clinical Trial. Frontiers in Pharmacology, 12, 614811. 10.3389/fphar.2021.614811

Lewis, D. A., & Hashimoto, T. (2007). Deciphering the disease process of schizophrenia: The contribution of cortical GABA neurons. International Review of Neurobiology, 78, 109–131. 10.1016/S0074-7742(06)78004-7

Lieberman, J. A., Girgis, R. R., Brucato, G., Moore, H., Provenzano, F., Kegeles, L., Javitt, D., Kantrowitz, J., Wall, M. M., Corcoran, C. M., Schobel, S. A., & Small, S. A. (2018). Hippocampal dysfunction in the pathophysiology of schizophrenia: A selective review and hypothesis for early detection and intervention. Molecular Psychiatry, 23(8), Article 8. 10.1038/mp.2017.249

Martin-Santos, R., Crippa, J. A., Batalla, A., Bhattacharyya, S., Atakan, Z., Borgwardt, S., Allen, P., Seal, M., Langohr, K., Farré, M., Zuardi, A. W., & McGuire, P. K. (2012). Acute effects of a single, oral dose of d9-tetrahydrocannabinol (THC) and cannabidiol (CBD) administration in healthy volunteers. Current Pharmaceutical Design, 18(32), 4966–4979. 10.2174/138161212802884780

McGuire, P., Robson, P., Cubala, W. J., Vasile, D., Morrison, P. D., Barron, R., Taylor, A., & Wright, S. (2017). Cannabidiol (CBD) as an Adjunctive Therapy in Schizophrenia: A Multicenter Randomized Controlled Trial. American Journal of Psychiatry, 175(3), 225–231. 10.1176/appi.ajp.2017.17030325

Modinos, G., Şimşek, F., Azis, M., Bossong, M., Bonoldi, I., Samson, C., Quinn, B., Perez, J., Broome, M. R., Zelaya, F., Lythgoe, D. J., Howes, O. D., Stone, J. M., Grace, A. A., Allen, P., & McGuire, P. (2018). Prefrontal GABA levels, hippocampal resting perfusion and the risk of psychosis. Neuropsychopharmacology, 43(13), 2652–2659. 10.1038/s41386-017-0004-6

Murphy, C. M., Christakou, A., Daly, E. M., Ecker, C., Giampietro, V., Brammer, M., Smith, A. B., Johnston, P., Robertson, D. M., MRC AIMS Consortium, Murphy, D. G., & Rubia, K. (2014). Abnormal Functional Activation and Maturation of Fronto-Striato-Temporal and Cerebellar Regions During Sustained Attention in Autism Spectrum Disorder. American Journal of Psychiatry, 171(10), 1107–1116. 10.1176/appi.ajp.2014.12030352

Nelson, E. A., Kraguljac, N. V., Maximo, J. O., Briend, F., Armstrong, W., Ver Hoef, L. W., Johnson, V., & Lahti, A. C. (2022). Hippocampal Dysconnectivity and Altered Glutamatergic Modulation of the Default Mode Network: A Combined Resting-State Connectivity and Magnetic Resonance Spectroscopy Study in Schizophrenia. Biological Psychiatry: Cognitive Neuroscience and Neuroimaging, 7(1), 108–118. 10.1016/j.bpsc.2020.04.014

O’Neill, A., Annibale, L., Blest-Hopley, G., Wilson, R., Giampietro, V., & Bhattacharyya, S. (2021). Cannabidiol modulation of hippocampal glutamate in early psychosis. Journal of Psychopharmacology, 026988112110011. 10.1177/02698811211001107

O’Neill, A., Wilson, R., Blest-Hopley, G., Annibale, L., Colizzi, M., Brammer, M., Giampietro, V., & Bhattacharyya, S. (2021). Normalization of mediotemporal and prefrontal activity, and mediotemporal-striatal connectivity, may underlie antipsychotic effects of cannabidiol in psychosis. Psychological Medicine, 51(4), 596–606. 10.1017/S0033291719003519

Pretzsch, C. M., Freyberg, J., Voinescu, B., Lythgoe, D., Horder, J., Mendez, M. A., Wichers, R., Ajram, L., Ivin, G., Heasman, M., Edden, R. A. E., Williams, S., Murphy, D. G. M., Daly, E., & McAlonan, G. M. (2019). Effects of cannabidiol on brain excitation and inhibition systems; a randomised placebo-controlled single dose trial during magnetic resonance spectroscopy in adults with and without autism spectrum disorder. Neuropsychopharmacology, 44(8), Article 8. 10.1038/s41386-019-0333-8

Provencher, S. W. (1993). Estimation of metabolite concentrations from localized in vivo proton NMR spectra. Magnetic Resonance in Medicine, 30(6), 672–679. 10.1002/mrm.1910300604

Rosenberg, E. C., Chamberland, S., Bazelot, M., Nebet, E. R., Wang, X., McKenzie, S., Jain, S., Greenhill, S., Wilson, M., Marley, N., Salah, A., Bailey, S., Patra, P. H., Rose, R., Chenouard, N., Sun, S. D., Jones, D., Buzsáki, G., Devinsky, O.,… Tsien, R. W. (2023). Cannabidiol modulates excitatory-inhibitory ratio to counter hippocampal hyperactivity. Neuron. 10.1016/j.neuron.2023.01.018

Salazar de Pablo, G., Radua, J., Pereira, J., Bonoldi, I., Arienti, V., Besana, F., Soardo, L., Cabras, A., Fortea, L., Catalan, A., Vaquerizo-Serrano, J., Coronelli, F., Kaur, S., Da Silva, J., Shin, J. I., Solmi, M., Brondino, N., Politi, P., McGuire, P., & Fusar-Poli, P. (2021). Probability of Transition to Psychosis in Individuals at Clinical High Risk. JAMA Psychiatry, 78(9), 1–9. 10.1001/jamapsychiatry.2021.0830

Santos-Silva, T., dos Santos Fabris, D., de Oliveira, C. L., Guimarães, F. S., & Gomes, F. V. (2024). Prefrontal and Hippocampal Parvalbumin Interneurons in Animal Models for Schizophrenia: A Systematic Review and Meta-analysis. Schizophrenia Bulletin, 50(1), 210–223. 10.1093/schbul/sbad123

Schobel, S. A., Chaudhury, N. H., Khan, U. A., Paniagua, B., Styner, M. A., Asllani, I., Inbar, B. P., Corcoran, C. M., Lieberman, J. A., Moore, H., & Small, S. A. (2013). Imaging patients with psychosis and a mouse model establishes a spreading pattern of hippocampal dysfunction and implicates glutamate as a driver. Neuron, 78(1), 81–93. 10.1016/j.neuron.2013.02.011

Schobel, S. A., Lewandowski, N. M., Corcoran, C. M., Moore, H., Brown, T., Malaspina, D., & Small, S. A. (2009). Differential targeting of the CA1 subfield of the hippocampal formation by schizophrenia and related psychotic disorders. 66(9), 938–946. 10.1001/archgenpsychiatry.2009.115

Smucny, J., Dienel, S. J., Lewis, D. A., & Carter, C. S. (2022). Mechanisms underlying dorsolateral prefrontal cortex contributions to cognitive dysfunction in schizophrenia. Neuropsychopharmacology, 47(1), 292–308. 10.1038/s41386-021-01089-0

Spielberger, C. D., Gonzalez-Reigosa, F., & Angel Martinez-Urrutia. (1971). DEVELOPMENT OF THE SPANISH EDITION OF THE STATE-TRAIT ANXIETY INVENTORY.

Talati, P., Rane, S., Skinner, J., Gore, J., & Heckers, S. (2015). Increased hippocampal blood volume and normal blood flow in schizophrenia. Psychiatry Research: Neuroimaging, 232(3), 219–225. 10.1016/j.pscychresns.2015.03.007

Uhlhaas, P. J. (2013). Dysconnectivity, large-scale networks and neuronal dynamics in schizophrenia. Current Opinion in Neurobiology, 23(2), 283–290. 10.1016/j.conb.2012.11.004

Uhlhaas, P. J., & Singer, W. (2010). Abnormal neural oscillations and synchrony in schizophrenia. Nature Reviews Neuroscience, 11(2), 100–113. 10.1038/nrn2774

Valli, I., Stone, J., Mechelli, A., Bhattacharyya, S., Raffin, M., Allen, P., Fusar-Poli, P., Lythgoe, D., O’Gorman, R., Seal, M., & McGuire, P. (2011). Altered Medial Temporal Activation Related to Local Glutamate Levels in Subjects with Prodromal Signs of Psychosis. Biological Psychiatry, 69(1), 97–99. 10.1016/j.biopsych.2010.08.033

Wilson, R., Bossong, M. G., Appiah-Kusi, E., Petros, N., Brammer, M., Perez, J., Allen, P., McGuire, P., & Bhattacharyya, S. (2019). Cannabidiol attenuates insular dysfunction during motivational salience processing in subjects at clinical high risk for psychosis. Translational Psychiatry, 9, 203. 10.1038/s41398-019-0534-2

Yung, A. R., Yuen, H. P., McGorry, P. D., Phillips, L. J., Kelly, D., Dell’Olio, M., Francey, S. M., Cosgrave, E. M., Killackey, E., Stanford, C., Godfrey, K., & Buckby, J. (2005). Mapping the onset of psychosis: The Comprehensive Assessment of At-Risk Mental States. The Australian and New Zealand Journal of Psychiatry, 39(11–12), 964–971. 10.1080/j.1440-1614.2005.01714.x

Zahid, U., Onwordi, E. C., Hedges, E. P., Wall, M. B., Modinos, G., Murray, R. M., & Egerton, A. (2023). Neurofunctional correlates of glutamate and GABA imbalance in psychosis: A systematic review. Neuroscience & Biobehavioral Reviews, 144, 105010. 10.1016/j.neubiorev.2022.105010

