## Supplemental Materia for "A single dose of cannabidiol modulates the coupling between hippocampal glutamate and learning-related prefrontal activation in individuals at Clinical High Risk of Psychosis"

**Supplementary Online Content**

### **eMethods**

#### ***Experimental procedure***

The procedure has been published in previous studies (Bhattacharyya et al., 2018; Davies et al., 2023a; Wilson et al., 2019).

In a parallel-group, double-blind, randomized, and placebo-controlled study, CHR participants were assigned to either a placebo group (CHR-PLB) or a CBD treatment group (CHR-CBD). Both participants and researchers collecting data were blinded to the treatment assignments. The randomization sequence, which ensured balanced allocation between the two groups using a blocking factor, was generated by STI Pharmaceuticals. A clinical trial pharmacist, who was not involved in other aspects of the study and remained unblinded, assigned participants to one of the two groups according to the randomization list. Once assigned, participants remained in their respective treatment arms. Allocation details were securely stored at the Maudsley hospital pharmacy.

Participants in the CHR-CBD group received a single 600 mg oral dose of CBD, while those in the CHR-PLB group were given a placebo capsule. The 600 mg/day dose was chosen based on previous evidence indicating that this dose range (600-800 mg/day) was as effective as antipsychotic medications (such as amisulpride) in reducing symptoms in patients with first-episode psychosis (Leweke et al., 2012). After receiving their respective treatments, participants underwent functional magnetic resonance imaging (fMRI) while performing a verbal paired-associate learning task that lasted approximately 13 minutes. MRI scans commenced three hours after administration of the study drug (CBD or placebo), based on prior research indicating peak CBD levels occur around that time following oral intake (Martin-Santos et al., 2012).

Healthy control (HC) participants were evaluated under identical conditions but did not receive any study drug. On the study day, all participants consumed a standardized light breakfast in front of the researchers and were instructed to abstain from cannabis for 96 hours, alcohol for at least 24 hours, and nicotine for 6 hours before scanning. Additionally, they were required to avoid any recreational drug use for two weeks prior to the study. Before scanning, a urine test was conducted to screen for illicit drug use. Three hours before the scan, participants in the CHR-CBD group took one cannabidiol capsule (600 mg, approximately 99.9% pure, sourced from THC-Pharm, Frankfurt, Germany), while participants in the CHR-PLB group took a visually identical placebo capsule filled with flour.

#### ***1H-MRS acquisition and quantification***

All MRI data was collected using a Signa HDx 3T MRI scanner (General Electric). A high-resolution inversion recovery image was also acquired. Full details of ^1^H-MRS acquisition parameters, preprocessing and analysis methods have been reported elsewhere (Davies et al., 2023a). In brief, we acquired ^1^H-MRS spectra using PRESS (Point RESolved Spectroscopy; TE = 30ms; TR = 3000ms; 96 averages) in the left hippocampus, following previously established methods (Egerton et al., 2014) using the standard GE PROBE (proton brain examination) sequence, which uses a standardised chemically selective suppression (CHESS) water suppression routine. For each metabolite spectrum, we also acquired unsuppressed water reference spectra (16 averages) as part of the standard acquisition for subsequent eddy current correction and water scaling. Auto-prescan was performed twice before each scan to optimise shimming and water suppression. The left hippocampal ^1^H-MRS voxel (20x20x15 mm; right-left, anterior-posterior, superior-inferior; positioned over the centre of the left hippocampus *(e****Figure 1***) for spectra acquisition was prescribed from a whole-brain 3D sagittal T1-weighted scan (TE=2.85ms; TR=6.98ms; TI=400ms; flip angle=11°, voxel size=1.0x1.0x1.2mm) using standardised protocols.

LCModel version 6.3-0A (Provencher, 1993) was used to analyse the acquired spectra, with a standard basis set of 16 metabolites (L-alanine, aspartate, creatine, phosphocreatine, GABA, glucose, glutamine, glutamate, glycerophosphocholine, glycine, myo-inositol, L-lactate, N-acetylaspartate, N-acetylaspartylglutamate, phosphocholine, and taurine) as detailed in the LCModel manual (<http://s-provencher.com/pub/LCModel/manual/manual.pdf>). Metabolite analyses were restricted to those meeting the quality control criteria of Cramér–Rao lower bounds ≤ 20% and signal to noise ratio ≥ 5. We corrected values of the water-scaled measure of glutamate for voxel tissue and cerebral spinal fluid (CSF) content using the formula Mcorr = M * (WM + 1.21GM + 1.55CSF) / (WM + GM), where M is the uncorrected metabolite value, and WM, GM and CSF are the white matter, grey matter and CSF fractions in the voxel, respectively, following previous work (Egerton et al., 2014) (assuming a CSF water concentration of 55,556 mol/m^3^ and the LCModel default brain water concentration of 35,880 mol/m^3^ (Gasparovic et al., 2006; Kreis et al., 1993)). For each participant, we determined these fractions from their structural T1 images using Statistical Parametric Mapping 8 software (SPM8), which was used to localise the spectroscopy voxel and subsequently segment into grey matter, white matter, and CSF.

#### ***fMRI processing and subject-level analysis details***

Instead of assuming a Gaussian population distribution of fMRI signals in the brain (an assumption that is difficult to test and found to be violated when tested (Brammer et al., 1997; Thirion et al., 2007a)), XBAM uses median instead of mean as a test statistic and permutation-based rather than normal theory-based approaches for statistical inference. As a result, it is less likely to suffer from bias caused by outliers (Hayasaka & Nichols, 2003). In this method, computation of the test statistic involves standardizing for individual differences in residual noise before embarking on second-level, multi-subject testing, using robust permutation-based methods, employing a mixed-effects approach to deal with the issue of non-normality. The use of a mixed effects approach helps address the inequality of individual residual variances by effectively “down weighting” responses with large residual variances. The significance of the resulting reweighted responses at the group level is then tested by data permutation to avoid assumptions of normality. Before analysis, the images were corrected for head motion (Brammer et al., 1997; Bullmore et al., 1999) and spatially smoothed with an 8.8mm full-width-at-half-maximum Gaussian filter (Thirion et al., 2007b). Slice timing correction was then applied, and the residual motion effects were regressed out from the time series using the estimated motion parameters before fitting a general linear model. The experimental design (encoding and recall condition) was convolved with 2 γ-variate functions, peaking at 4 and 8 seconds to allow for variability in hemodynamic delay to model the BOLD response. To estimate the BOLD response, a constrained BOLD effects model was used, and the best fit was computed between the weighted sum of convolutions and the change over time at each voxel, which helped to reduce the possibility that increased mathematically plausible but decreased physiologically implausible results caused by the model fitting procedure. This model was then fitted to the time series at each voxel using least-squares fitting. Following this, the sum of squares (SSQ) ratio statistic, which represents the ratio of the model component to the residual sum of squares, was estimated for the encoding and correct recall conditions relative to the baseline during which a blank screen is shown to participants (encoding vs baseline; recall vs baseline)

The SSQ ratio maps of each condition for each individual were then transformed into standard stereotactic space (Talairach space).

### **eFigures**

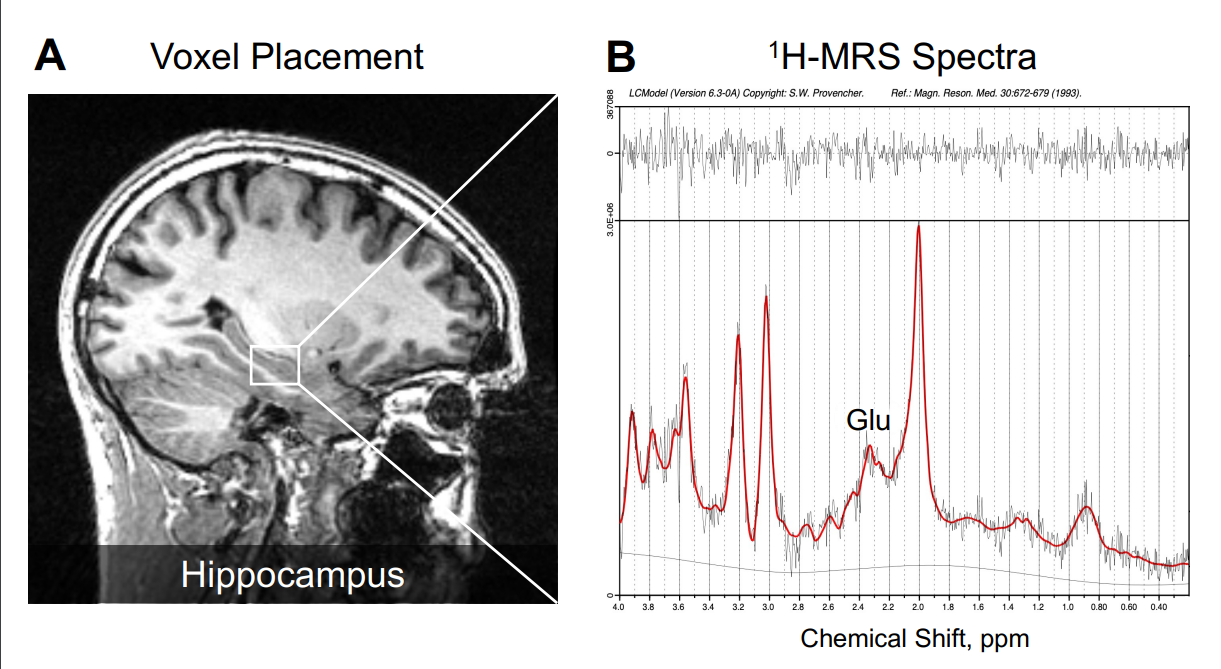

#### eFigure 1. Example 1H-MRS voxel positioning and spectra in left hippocampus.

**eFigure 1.** In panel A, an illustrative example of voxel placement in the left hippocampus is indicated by the box. In panel B, 1H-MRS spectrum obtained (black line) from the voxel in A and the overlay of the spectral fit (red line). All spectra were analysed with LCModel version 6.3-0A (S.W. Provencher). Glu indicates glutamate and ppm, parts per million (Davies et al., 2023a).

#### eFigure 2: pre-defined anatomical mask

*:*
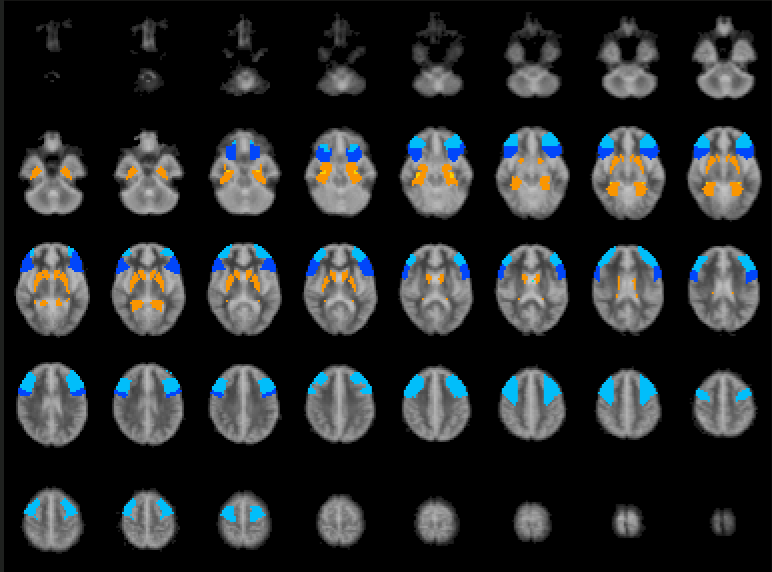

eFigure 2. The pre-defined anatomical mask used for region of interest (ROI) analysis included the bilateral hippocampus, parahippocampal gyrus, striatum, and inferior and middle frontal gyrus, shown here overlaid on the Talairach template.

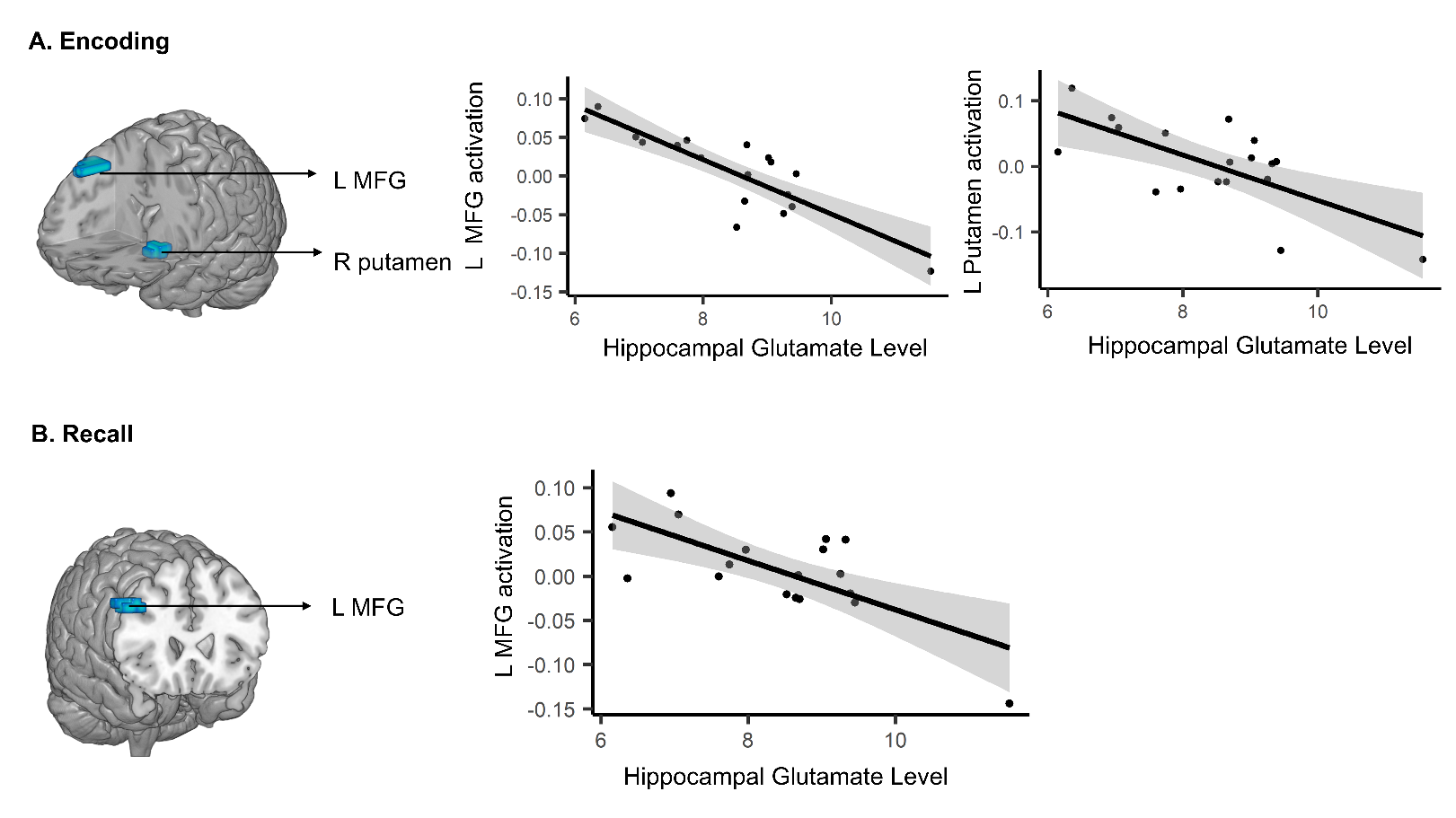

#### eFigure 3. Association between hippocampal glutamate levels and brain activation in healthy controls

Figure 3: Clusters of brain activation (as indexed by the sum of squares ratio, SSQ) showing significant correlations with left hippocampal glutamate levels in healthy controls. The brain images display significant clusters identified in the glutamate x BOLD x group interaction analysis, and the scatterplots show the relationship between left hippocampal glutamate levels and the SSQ value extracted from the corresponding cluster. A. Encoding: The left line plot shows a cluster in the left middle frontal cortex (peak Talairach coordinates: X = -29, Y = 22, Z = 46; Cluster size = 24 voxels; p = 0.0001) where a negative correlation was found between regional activation and left hippocampal glutamate levels during the encoding condition. The right line plots shows a cluster in the right putamen (peak Talairach coordinates: X = 22, Y = 7, Z = -10; Cluster size = 10 voxels; p = 0.0029) where there was also a negative correlation between regional activation and left hippocampal glutamate levels during encoding. B. Recall: The line plot shows a cluster in the left middle frontal cortex (peak Talairach coordinates: X = -43, Y = 22, Z = 30; Cluster size = 24 voxels; p = 0.0001) where a negative correlation was found between regional activation and left hippocampal glutamate levels during the recall sessions.

Bhattacharyya, S., Wilson, R., Appiah-Kusi, E., O’Neill, A., Brammer, M., Perez, J., Murray, R., Allen, P., Bossong, M. G., & McGuire, P. (2018). Effect of Cannabidiol on Medial Temporal, Midbrain, and Striatal Dysfunction in People at Clinical High Risk of Psychosis: A Randomized Clinical Trial. *JAMA Psychiatry*, *75*(11), 1107–1117. https://doi.org/10.1001/jamapsychiatry.2018.2309

Brammer, M. J., Bullmore, E. T., Simmons, A., Williams, S. C. R., Grasby, P. M., Howard, R. J., Woodruff, P. W. R., & Rabe-Hesketh, S. (1997). Generic brain activation mapping in functional magnetic resonance imaging: A nonparametric approach. *Magnetic Resonance Imaging*, *15*(7), 763–770. https://doi.org/10.1016/S0730-725X(97)00135-5

Bullmore, E. T., Suckling, J., Overmeyer, S., Rabe-Hesketh, S., Taylor, E., & Brammer, M. J. (1999). Global, voxel, and cluster tests, by theory and permutation, for a difference between two groups of structural MR images of the brain. *IEEE Transactions on Medical Imaging*, *18*(1), 32–42. IEEE Transactions on Medical Imaging. https://doi.org/10.1109/42.750253

Davies, C., Bossong, M. G., Martins, D., Wilson, R., Appiah-Kusi, E., Blest-Hopley, G., Allen, P., Zelaya, F., Lythgoe, D. J., Brammer, M., Perez, J., McGuire, P., & Bhattacharyya, S. (2023a). Hippocampal Glutamate, Resting Perfusion and the Effects of Cannabidiol in Psychosis Risk. *Schizophrenia Bulletin Open*, *4*(1), sgad022. https://doi.org/10.1093/schizbullopen/sgad022

Egerton, A., Stone, J. M., Chaddock, C. A., Barker, G. J., Bonoldi, I., Howard, R. M., Merritt, K., Allen, P., Howes, O. D., Murray, R. M., McLean, M. A., Lythgoe, D. J., O’Gorman, R. L., & McGuire, P. K. (2014). Relationship Between Brain Glutamate Levels and Clinical Outcome in Individuals at Ultra High Risk of Psychosis. *Neuropsychopharmacology*, *39*(12), Article 12. https://doi.org/10.1038/npp.2014.143

Gasparovic, C., Song, T., Devier, D., Bockholt, H. J., Caprihan, A., Mullins, P. G., Posse, S., Jung, R. E., & Morrison, L. A. (2006). Use of tissue water as a concentration reference for proton spectroscopic imaging. *Magnetic Resonance in Medicine*, *55*(6), 1219–1226. https://doi.org/10.1002/mrm.20901

Hayasaka, S., & Nichols, T. E. (2003). Validating cluster size inference: Random field and permutation methods. *NeuroImage*, *20*(4), 2343–2356. https://doi.org/10.1016/j.neuroimage.2003.08.003

Kreis, R., Ernst, T., & Ross, B. D. (1993). Absolute Quantitation of Water and Metabolites in the Human Brain. II. Metabolite Concentrations. *Journal of Magnetic Resonance, Series B*, *102*(1), 9–19. https://doi.org/10.1006/jmrb.1993.1056

Leweke, F. M., Piomelli, D., Pahlisch, F., Muhl, D., Gerth, C. W., Hoyer, C., Klosterkötter, J., Hellmich, M., & Koethe, D. (2012). Cannabidiol enhances anandamide signaling and alleviates psychotic symptoms of schizophrenia. *Translational Psychiatry*, *2*(3), e94–e94. https://doi.org/10.1038/tp.2012.15

Martin-Santos, R., Crippa, J. A., Batalla, A., Bhattacharyya, S., Atakan, Z., Borgwardt, S., Allen, P., Seal, M., Langohr, K., Farré, M., Zuardi, A. W., & McGuire, P. K. (2012). Acute effects of a single, oral dose of d9-tetrahydrocannabinol (THC) and cannabidiol (CBD) administration in healthy volunteers. *Current Pharmaceutical Design*, *18*(32), 4966–4979. https://doi.org/10.2174/138161212802884780

Provencher, S. W. (1993). Estimation of metabolite concentrations from localized in vivo proton NMR spectra. *Magnetic Resonance in Medicine*, *30*(6), 672–679. https://doi.org/10.1002/mrm.1910300604

Thirion, B., Pinel, P., Mériaux, S., Roche, A., Dehaene, S., & Poline, J.-B. (2007a). Analysis of a large fMRI cohort: Statistical and methodological issues for group analyses. *NeuroImage*, *35*(1), 105–120. https://doi.org/10.1016/j.neuroimage.2006.11.054

Thirion, B., Pinel, P., Mériaux, S., Roche, A., Dehaene, S., & Poline, J.-B. (2007b). Analysis of a large fMRI cohort: Statistical and methodological issues for group analyses. *NeuroImage*, *35*(1), 105–120. https://doi.org/10.1016/j.neuroimage.2006.11.054

Wilson, R., Bossong, M. G., Appiah-Kusi, E., Petros, N., Brammer, M., Perez, J., Allen, P., McGuire, P., & Bhattacharyya, S. (2019). Cannabidiol attenuates insular dysfunction during motivational salience processing in subjects at clinical high risk for psychosis. *Translational Psychiatry*, *9*, 203. https://doi.org/10.1038/s41398-019-0534-2
